# The Relationship of Antepartum Fetal Heart Rate Patterns to Adverse Pregnancy Outcomes

**DOI:** 10.1101/2024.11.16.24317432

**Authors:** William Cooke, Beth Albert, Manu Vatish, Gabriel Davis Jones

**Affiliations:** Nuffield Department of Women’s & Reproductive Health, University of Oxford, Women’s Centre, John Radcliffe Hospital, Oxford, OX3 9DU, United Kingdom; The Alan Turing Institute, London, UK

## Abstract

**Introduction:** Antepartum fetal heart rate (FHR) patterns are routinely assessed to evaluate fetal wellbeing. Despite their clinical use, the relationship between specific FHR patterns and adverse pregnancy outcomes (APOs) remains unclear. This study aims to investigate the association between antepartum FHR patterns and APOs to improve fetal risk assessment.

**Methods:** In this retrospective case–control study, we extracted raw antepartum FHR traces from singleton pregnancies between 27^+0^ and 41^+6^ weeks’ gestation recorded at Oxford University Hospitals from January 1991 to February 2024. Adverse outcomes included acidaemia, stillbirth, asphyxia, extended neonatal care unit (NCU) admission, hypoxic–ischaemic encephalopathy (HIE), low Apgar scores and neonatal resuscitation at delivery. After applying inclusion and exclusion criteria, 938 FHR traces with APOs were matched using propensity score matching with 938 traces from normal pregnancy outcomes (NPOs), controlling for gestational age, fetal sex, maternal BMI, maternal age, parity, and trace duration. FHR patterns were extracted using a validated automated algorithm and analysed statistically.

**Results:** The APO cohort showed significantly higher basal heart rates (BHR), fewer accelerations, more decelerations, lower short-term variability (STV), and spent a greater proportion of the trace in periods of low variation compared to the NPO cohort (*p* ***<* 0.001**). Logistic regression identified prolonged periods of low variation (odds ratio [OR] = 1.92, 95% CI 1.60–2.30, *p* ***<* 0.001**), increased decelerations (OR = 1.40, 95% CI 1.22–1.60, *p* ***<* 0.001**), reduced accelerations (OR = 0.66, 95% CI 0.55–0.78, *p* ***<* 0.001**), elevated BHR (OR = 0.70, 95% CI 0.61–0.80, *p* ***<* 0.001**), and decreased STV (OR = 0.72, 95% CI 0.57–0.91, *p* **= 0.006**) as significant predictors of APOs.

**Conclusions:** Specific antepartum FHR patterns are significantly associated with adverse pregnancy outcomes. Detailed analysis of these patterns can enhance fetal risk assessment and inform clinical decision-making. Adoption of standardised interpretation criteria for antepartum FHR monitoring may improve perinatal outcomes.

## 1 Introduction

The fetal heart rate (FHR) is primarily controlled by the central and autonomic nervous systems. Patterns within the FHR are therefore used to evaluate brain health and fetal wellbeing. The cardinal antepartum FHR patterns include basal heart rate (BHR), accelerations, decelerations, deceleration magnitude, short-term variability (STV), episodes of high variation and episodes of low variation.[1] The BHR is the average FHR measured over a period, excluding significant fluctuations. Accelerations are transient increases in heart rate, while decelerations are temporary decreases. The deceleration magnitude refers to the depth and duration. STV captures the beat-to-beat fluctuations in the FHR. Episodes of high variation involve greater oscillations in the FHR, whereas episodes of low variation are characterised by a more stable and less fluctuating FHR.

Despite the routine use of these patterns in computerised antenatal fetal monitoring (cardiotocography)[1] and their incorporation into clinical guidelines,[2] the relationship between these characteristics and normal or adverse pregnancy outcomes is not yet fully understood in the antepartum period. Established guidelines exist for interpreting FHR patterns during labour, where the fetus is exposed to intermittent hypoxic stress due to uterine contractions, variations in placental blood flow and changes in maternal oxygenation.[3] However, these stressors do not generally exist in the antepartum. Thus, the connection between antepartum FHR patterns and outcomes such as stillbirth or acidaemia remains less clear.

Despite advancements in computerised analysis, there remains limited understanding of the relative correlation between each component of the antepartum FHR pattern and adverse fetal outcomes. This gap underscores the need for further research to understand the value of FHR monitoring in the antepartum period, with the goal of enhancing risk stratification and improving pregnancy outcomes.

We conducted a comprehensive evaluation of antepartum FHR patterns using computerised antepartum FHR monitoring. This retrospective case-control study, using data from Oxford University Hospitals, UK, aimed to assess the utility of these patterns and their relative importance to each other in differentiating between adverse and normal pregnancy outcome cohorts. The objective was to enhance our understanding of fetal physiology, fetal risk assessment and improve the predictive accuracy of antepartum FHR monitoring.

## 2 Methods

### 2.1 Data processing, cohort development, and extraction of fetal heart rate patterns

We extracted raw digital antepartum FHR traces from the Oxford University Hospitals maternity database at the John Radcliffe Hospital (Oxford, United Kingdom) for the period between 1 January 1991 and 1 February 2024. Traces were obtained from singleton pregnancies between 27^+0^ and 41^+6^ gestational weeks, with available associated clinical outcome data for both the mother and baby. Traces with more than 30% signal loss or a duration of less than 30 minutes were excluded from the analysis.

The adverse pregnancy outcomes (APOs) considered in this study were acidaemia, stillbirth, asphyxia, extended neonatal care unit (NCU) admission, hypoxic–ischaemic encephalopathy (HIE), low Apgar score, and neonatal resuscitation at delivery (requiring either cardiac massage and/or THAM/bicarbonate administration). Supplementary Table 1 details definitions and justifications for these adverse outcomes. The definitions and reference values used were in alignment with those used by NICE, FIGO, and ACOG for pH thresholds for acidaemia, Apgar scoring, stillbirth definition, and neonatal resuscitation. Only cases involving APOs that underwent elective or emergency caesarean section within 24 hours of the antepartum FHR trace were included, as outcomes *>*24 hours after antepartum CTG were considered too temporally distant to be meaningfully correlated.[4]

To establish the normal pregnancy outcome (NPO) cohort, specific inclusion and exclusion criteria were applied. Pregnancies were included if they met the following criteria: a liveborn singleton baby, gestational age at delivery between 37^+0^ and 41^+6^ weeks, maternal age at booking between 18 and 40 years, birthweight at or above the 10th centile for gestational age, labour duration of less than 24 hours, Apgar scores of *>*4 at 1 minute and *>*7 at 5 minutes, and normal umbilical venous or arterial pH values. For caesarean deliveries without labour, an arterial pH *>*7.13 and base deficit *<* 10.0 were required, while for deliveries following labour, an arterial pH *>*7.05 and base deficit *<*14.0 were considered.

Data were excluded from the NPO cohort if any of the following conditions were met: neonatal death within 3 months, delivery by emergency caesarean section, any requirement for neonatal resuscitation, NCU admission after birth, neonatal cooling, hypertensive disorders of pregnancy such as pre-eclampsia, or clinically suspected infection or sepsis.

Antepartum FHR traces from the NPO cohort were matched with those from the APO cohort using one-to-one propensity score matching (PSM) and had their FHR patterns extracted using a clinically validated automated algorithm.[1, 5] PSM is a statistical technique used to reduce bias by matching subjects from different groups based on similar propensity scores, which are calculated using a set of observed factors. This matching controlled for six factors: gestational age at FHR monitoring, fetal sex, maternal BMI at booking, maternal age at delivery, parity, and duration of the FHR trace in minutes. By controlling for these variables, each case in the NPO cohort was paired with a directly comparable counterpart in the APO cohort, thereby minimising potential bias.[1]

Propensity scores were estimated using logistic regression, with outcome assignment coded as 1 for the APO cohort and 0 for the NPO cohort.[6] Interaction terms were incorporated to assess whether the effect of one covariate on the outcome varied depending on another covariate. Matching was performed using a Nearest Neighbours model, implemented with a ball tree algorithm. A caliper of 0.05 was applied to improve match precision and quality. Matching quality was evaluated through multiple metrics to ensure balanced cohorts. Covariate balance was assessed using Standardised Mean Differences (SMDs), with an SMD below 0.10 considered acceptable. Histograms, bar charts, area under the receiver-operator curve (AU-ROC), and Brier Score were employed to evaluate the effectiveness of the matching process.[6]

### 2.2 Statistical analysis

Discrete variables are presented as counts with interquartile ranges (IQRs) and percentages, while continuous variables are summarised using medians and interquartile ranges (IQRs). For comparisons, categorical variables were analysed using the Chi-square test, and continuous variables were assessed using the Mann-Whitney U test, with statistical significance set at *p <* 0.05. Cohen’s d was calculated to evaluate effect size. Logistic regression was performed on scaled FHR pattern values to evaluate the association between FHR patterns and adverse outcomes, with results expressed as coefficients, odds ratios (ORs), and their respective 95% confidence intervals (CIs). All analyses were conducted using Python (version 3.9.17), employing the Pandas (version 1.5.3), NumPy (version 1.23.5), Matplotlib (version 3.7.1), SciPy (version 1.10.1), and Statsmodels (version 0.13.5) libraries.

## 3 Results

A total of 938 fetal heart rate (FHR) traces with adverse pregnancy outcomes (APO) were matched with 938 traces from normal pregnancy outcomes (NPO), resulting in 1,876 FHR records. Propensity score matching (PSM) created well-balanced cohorts with an AU-ROC of 0.81 (95% CI 0.78–0.83) and a Brier Score of 0.03. The median gestational age when the FHR trace was performed was 34.5 weeks (IQR 30.0–38.0) for both the APO and NPO cohorts (SMD = 0.00). The median duration of the FHR trace was 60.0 minutes (IQR 55.0–60.0) for both cohorts. The sex ratio was 54.3% and 54.6% males in the NPO and APO cohorts, respectively, with females accounting for 45.7% and 45.4% (SMD = 0.00). The mean maternal BMI at booking was 26.3 kg/m^2^ (IQR 23.0–30.7) for the APO cohort and 26.1 kg/m^2^ (IQR 22.9–30.5) for the NPO cohort (SMD = 0.08). The median maternal age at delivery was 31.0 years (IQR 27.0–35.0) in both cohorts (SMD = 0.00). Median gravidity was 1 (IQR 0–2) and parity was 0 (IQR 0–1) for both cohorts (SMD = 0.00).

Labour and delivery outcomes differed significantly between cohorts. In the APO cohort, 16.9% underwent induced labour compared to 29.9% in the NPO cohort, with spontaneous labour accounting for 11.2% in APO and 54.2% in NPO. Delivery methods also varied, as the APO cohort underwent emergency (28.2%) or elective (71.8%) caesarean sections, whereas spontaneous deliveries were predominant in the NPO cohort (62.9%).

### 3.1 Newborn characteristics

Newborn characteristics showed notable differences between cohorts. Apgar scores at 1 minute were significantly lower in the APO cohort, with a median of 3.0 (IQR 2.0–5.0), compared to 10.0 (IQR 9.0–10.0) in the NPO cohort. At 5 minutes, the median Apgar score in the APO cohort improved to 8.0 (IQR 6.0–10.0) compared to 10.0 (IQR 10.0–10.0) in the NPO cohort. The median umbilical artery pH was 7.21 (IQR 7.10–7.27) in the APO cohort and 7.25 (IQR 7.20–7.30) in the NPO cohort. Venous pH levels were 7.26 (IQR 7.16–7.32) in APO and 7.33 (IQR 7.28–7.36) in NPO. Characteristics of mothers and newborns are presented in Supplementary Table 2.

### 3.2 Comparative analysis of FHR patterns

In comparing FHR patterns between APO and NPO cohorts, several significant differences were observed (Table 1). The BHR was marginally higher in the APO cohort, with a median of 142.0 bpm (IQR 135.0–150.0) compared to 140.0 bpm (IQR 134.0–147.0) in the NPO cohort (Cohen’s d = -0.21, *p <* 0.001). The number of accelerations was significantly lower in the APO group, with a median of 3.0 (IQR 0.0–8.0) compared to 9.0 (IQR 5.0–15.0) in the NPO group (Cohen’s d = 0.81, *p <* 0.001). Decelerations were similar between the cohorts, with both having a median of 1.0 (IQR 0.0–2.0), though the difference was statistically significant (Cohen’s d = -0.12, *p <* 0.01). The largest deceleration magnitude was higher in the APO cohort, with a median of 16.0 lost beats (IQR 9.0–36.0) compared to 10.0 (IQR 8.0–21.2) in the NPO cohort (Cohen’s d = -0.14, *p <* 0.001). STV was significantly lower in the APO group, with a median of 5.5 (IQR 4.2–7.5) compared to 7.9 (IQR 6.4–9.8) in the NPO group (Cohen’s d = 0.81, *p <* 0.001).

**Table 1.** Comparison of antepartum fetal heart rate (FHR) patterns between normal pregnancy outcomes (NPO) and adverse pregnancy outcomes (APO). Data are presented as median (interquartile range). Effect size and p-values indicate the magnitude and significance of differences. Abbreviations: LDM, largest deceleration magnitude; STV, short-term variability; High Variation and Low Variation represent the proportion of the total length of the FHR trace.

| FHR Pattern | NPO | APO | Effect size | p-value |
| --- | --- | --- | --- | --- |
| BHR | 140.0 (134.0–147.0) | 142.0 (135.0–150.0) | -0.21 | < 0.001 |
| Accelerations | 9.0 (5.0–15.0) | 3.0 (0.0–8.0) | 0.81 | < 0.001 |
| Decelerations | 1.0 (0.0–2.0) | 1.0 (0.0–2.0) | -0.12 | < 0.01 |
| LDM | 10.0 (8.0–21.2) | 16.0 (9.0–36.0) | -0.14 | < 0.001 |
| STV | 7.9 (6.4–9.8) | 5.5 (4.2–7.5) | 0.81 | < 0.001 |
| High Variation | 0.3 (0.1–0.6) | 0.1 (0.0–0.3) | 0.78 | < 0.001 |
| Low Variation | 0.1 (0.0–0.3) | 0.4 (0.2–0.7) | -0.88 | < 0.001 |

### 3.3 Regression analysis

Logistic regression evaluating the association between FHR patterns and APO revealed significant predictors (Table 2). The basal heart rate was significantly associated with APO, showing a negative coefficient of -0.36 (95% CI -0.49—0.22) and an odds ratio (OR) of 0.70 (95% CI 0.61–0.80, *p <* 0.001). Accelerations were negatively associated, with a coefficient of -0.42 (95% CI -0.60—0.24) and an OR of 0.66 (95% CI 0.55–0.78, *p <* 0.001). Decelerations were positively associated with APO, with a coefficient of 0.34 (95% CI 0.20–0.47) and an OR of 1.40 (95% CI 1.22–1.60, *p <* 0.001). STV was negatively associated with APO, with a coefficient of -0.33 (95% CI -0.57—0.09) and an OR of 0.72 (95% CI 0.57–0.91, *p* = 0.006).

**Table 2.** Odds ratios (95% confidence intervals) for fetal heart rate (FHR) patterns associated with adverse pregnancy outcomes (APO). The direction of association is indicated by arrows: ↑ increased association, ↓ decreased association, – no association. P-values denote the statistical significance of the associations. Abbreviations: LDM, largest deceleration magnitude; STV, short-term variability; High Variation and Low Variation represent the proportion of the total length of the FHR trace.

| FHR Pattern | Odds Ratio (95% CI) | Association with APO | p-value |
| --- | --- | --- | --- |
| BHR | 0.70 (0.61–0.80) | ↓ | $< 0.001$ |
| Accelerations | 0.66 (0.55–0.78) | ↓ | $< 0.001$ |
| Decelerations | 1.40 (1.22–1.60) | ↑ | $< 0.001$ |
| LDM | 1.05 (0.93–1.18) | $-$ | 0.471 |
| STV | 0.72 (0.57–0.91) | ↓ | $< 0.01$ |
| High Variation | 1.01 (0.82–1.24) | $-$ | 0.941 |
| Low Variation | 1.92 (1.60–2.30) | ↑ | $< 0.001$ |

## 4 Discussion

In this retrospective case-control study involving 1,876 antepartum fetal heart rate (FHR) traces, we identified significant differences in FHR patterns between pregnancies with adverse pregnancy outcomes (APOs) and those with normal pregnancy outcomes (NPOs). The APO cohort exhibited higher basal heart rates (BHR), fewer accelerations, more decelerations, lower short-term variability (STV), and spent a greater proportion of the FHR trace in periods of low variation compared to the NPO cohort. These differences were consistent across various APOs, including acidaemia, stillbirth, asphyxia, extended neonatal care unit (NCU) admission, hypoxic–ischaemic encephalopathy (HIE), low Apgar scores, and neonatal resuscitation at delivery. Sub-group analyses revealed these patterns were evident and generally consistent in both preterm and term pregnancies.

Logistic regression analysis revealed the proportion of the FHR trace spent in periods of low variation as the most significant predictor of APOs. With an odds ratio (OR) of 1.92 (95% CI 1.60–2.30, *p <* 0.001), this indicates that for each unit increase, the odds of experiencing an APO nearly double. This underscores the critical importance of prolonged low variability periods as a marker of fetal compromise. Decelerations were the next most important pattern, showing a positive association with APOs (OR = 1.40, 95% CI 1.22–1.60, *p <* 0.001). Increased frequency of decelerations suggests intermittent disruptions in fetal oxygenation, heightening the risk of adverse outcomes.[7]

Accelerations and higher BHR were both inversely associated with APOs (i.e., associated with a NPO). Accelerations had an OR of 0.66 (95% CI 0.55–0.78, *p <* 0.001), indicating that a higher number of accelerations significantly reduces the odds of an APO. Similarly, BHR had an OR of 0.70 (95% CI 0.61–0.80, *p <* 0.001), suggesting that higher basal rates are somewhat protective, possibly reflecting an appropriate fetal response to stress. STV showed a significant negative association with APOs (OR = 0.72, 95% CI 0.57–0.91, *p* = 0.006), but its effect size was smaller compared to high variation, decelerations, accelerations, and BHR. Patterns such as largest deceleration magnitude (LDM) and the proportion of time spent in high variation were not significantly associated with APOs (*p >* 0.05), suggesting they are less critical in predicting adverse outcomes or confirming normality.

The elevated BHR observed in the APO cohort may indicate fetal tachycardia, which can be a response to hypoxia or infection.[8, 9] Higher BHR could reflect a compensatory mechanism to maintain oxygen delivery to vital organs when fetal oxygenation is compromised. The significant reduction in accelerations suggests diminished fetal reactivity, possibly due to central nervous system depression associated with hypoxia or acidosis. Accelerations are typically indicative of fetal wellbeing, reflecting an intact autonomic nervous system and adequate oxygenation.[10]

The increase in decelerations in the APO group aligns with the understanding that decelerations can signify transient or sustained disruptions in fetal oxygenation.[11] Although the differences in deceleration frequency were modest, their presence in conjunction with other abnormal FHR patterns strengthens the concern for fetal compromise. The lower STV in the APO cohort is particularly noteworthy. STV reflects the interplay between sympathetic and parasympathetic nervous systems; decreased variability may indicate impaired autonomic control due to hypoxia or acid-base disturbances.[12]

The greater proportion of time spent in periods of low variation further emphasizes the potential for compromised fetal wellbeing in the APO group. Periods of low variation may represent sustained fetal quiescence or reduced responsiveness, which can be concerning when prolonged. These findings collectively suggest that specific antepartum FHR patterns are associated with an increased risk of adverse outcomes and could serve as early indicators of fetal distress.

These results are consistent with previous studies linking abnormal FHR patterns to adverse perinatal outcomes. Research has demonstrated that decreased accelerations and variability are associated with fetal acidaemia and hypoxia.[13, 14] For instance, reduced FHR variability and absent accelerations have been shown to associate with fetal compromise.[1, 15, 16] However, much of the existing literature has focused on intrapartum FHR monitoring, with well-established guidelines for interpretation during labour.[3, 17] Our study contributes to the limited body of research specifically examining antepartum FHR patterns. By identifying clear associations between antepartum FHR characteristics and a range of APOs, our findings support the utility of these patterns in prenatal risk assessment.

A key strength of this study is the large sample size and the use of propensity score matching to create well-balanced cohorts, minimising confounding variables such as gestational age, fetal sex, maternal BMI, maternal age, parity, and FHR trace duration. This methodological approach enhances the reliability of our comparisons between the APO and NPO groups. Nevertheless, the study has limitations. Being retrospective, it is subject to inherent biases related to data collection and documentation. We have sought to reduce this bias by constraining the interval between the FHR trace being acquired and caesarean section to 24 hours. The reliance on data from a single institution may limit the generalisability of the findings to other populations or settings with different patient demographics or clinical practices. We also acknowledge that while associations were identified, causality cannot be definitively established from this observational study.

The findings from this study offer important clinical insights into the use of antepartum FHR monitoring as a predictive tool for adverse pregnancy outcomes. Reduced accelerations, elevated basal heart rate, decreased short-term variability, and prolonged periods of low variability were found to be strong indicators of fetal compromise, reinforcing the need for vigilant monitoring in high-risk pregnancies. Clinically, these patterns can guide timely interventions, such as increasing surveillance or considering early delivery, to mitigate risks to both the fetus and the mother. The findings also highlight the potential for developing standardised guidelines for interpreting antepartum FHR patterns, analogous to those used during labour. Incorporating computerised analysis of FHR patterns into routine antepartum care could facilitate more objective assessments and reduce inter-observer variability. Ultimately, this could contribute to improved perinatal outcomes through timely and appropriate clinical responses.

Further research is warranted to validate these findings in prospective studies and diverse populations. Investigating the underlying pathophysiological mechanisms connecting specific FHR patterns to adverse pregnancy outcomes could provide deeper insights into fetal responses to stress. Additionally, integrating advanced computational techniques, such as machine learning algorithms, may enhance the predictive accuracy of FHR analyses. Future studies should also explore the cost-effectiveness and feasibility of implementing standardised antepartum FHR monitoring protocols in clinical practice.

## 5 Conclusions

This study demonstrates significant associations between specific antepartum FHR patterns and adverse pregnancy outcomes. Elevated basal heart rates, reduced accelerations and variability, increased decelerations, and prolonged periods of low variation were all linked to higher rates of APOs. These findings suggest detailed analysis of antepartum FHR patterns can enhance fetal risk assessment and potentially improve clinical outcomes. Adoption of standardised interpretation criteria for antepartum FHR monitoring may be beneficial in guiding clinical decision-making and warrants further investigation.

## Data Availability

The authors acknowledge the importance of data transparency and the potential value of data sharing in advancing scientific research. However, due to the identifiable and sensitive nature of the data used in this study, which includes detailed fetal heart rate traces potentially linked to individual patient outcomes, we are unable to make the dataset publicly available. The data contains protected health information and is subject to strict confidentiality constraints to safeguard the privacy of individuals. Consequently, the ethical and legal restrictions prevent the sharing of the dataset.

## Ethics

This study was approved by the Ethics Committee in the Joint Research Office, Research and Development Department, Oxford University Hospitals NHS Trust: 13/SC/0153.

## Acknowledgements

The authors wish to thank Mr Pawel Szafranski and Mr James Bland for their support in providing data acquisition and management services.

