## Supplementary Information for "The Relationship of Antepartum Fetal Heart Rate Patterns to Adverse Pregnancy Outcomes"

| Adverse Outcome | Definition | Justification |
|---|---|---|
| **Acidaemia** | Two sets of acidaemia values are used: Babies delivered by CS without labour: arterial pH <7.13 AND arterial BD >10.0; Babies who experienced labour (regardless of delivery method): arterial pH <7.05 and arterial BD >14.0. | Fetal acidaemia indicates a significant imbalance of pH levels in the fetus/neonate's blood, typically signifying a lack of oxygen. This condition can result in immediate threats to the infant's survival and long-term neurological damage if not promptly identified and treated.[1,2] |
| **Birth asphyxia** | Low Apgar score(s) + Acidaemia *Low Apgar score(s) are defined below* | Birth asphyxia occurs when there is an insufficient supply of oxygen to the baby before, during, or shortly after birth, leading to hypoxia and acidosis. This condition can cause organ failure and severe neurological issues, requiring immediate medical intervention to prevent lifelong disability or death.[3,4] |
| **Extended Neonatal Care Unit (NCU) admission** | Neonates born at or after $37^{+0}$ gestational weeks who were admitted for at least 7 days to NCU. | A prolonged stay in a special care unit indicates significant health issues requiring close medical attention, such as respiratory distress syndrome or severe infections. The need for extended NCU care can reflect the severity of the infant's condition, impacting their development and long-term health.[5,6] |
| **Hypoxic ischaemic encephalopathy (HIE)** | Diagnosed by treating clinical team (neonatal/paediatrics). | HIE is severe brain damage resulting from a lack of oxygen and blood flow to the brain, leading to immediate consequences like seizures and death, and long-term debilitating effects on developmental and cognitive abilities. Early diagnosis and intervention are crucial to mitigate the effects of HIE and improve the infant's prognosis.[7] |
| **Low Apgar score(s)** | Apgar score <4 at 1 minute; Apgar score <7 at 5 minutes. | The Apgar score assesses a newborn's heart rate, reflexes, muscle tone, skin colour, and respiration. A low Apgar score requires immediate medical intervention and potentially resuscitation, serving as a proxy for neonatal well-being and future health.[8,9] |
| **Neonatal resuscitation** | Either cardiac massage, sodium bicarbonate or and tris-hydroxymethyl-aminomethane (THAM) used following delivery of the fetus. | Neonatal resuscitation is an emergency procedure used to restore normal heart rate and breathing in a newborn experiencing life-threatening conditions such as severe acidosis, respiratory distress, or cardiac arrest. Immediate resuscitation is critical to prevent organ damage, neurological injury, or death. Interventions such as cardiac massage, sodium bicarbonate to counteract metabolic acidosis, and THAM to buffer blood pH are essential in stabilizing the infant.[10-14] |
| **Stillbirth** | Antepartum or intrapartum stillbirth, as diagnosed by treating clinical team. | Stillbirth, occurring before or during labour, or death within 7 days of delivery is a severe outcome with profound impact. Identifying risk factors early can |

lead to interventions that may save lives, making it a crucial high-risk condition for healthcare providers to monitor and manage through fetal monitoring and timely intervention.[15,16]

**Supplementary Table 1: Definitions of adverse outcomes utilised for inclusion in the adverse pregnancy outcome cohort.** Each outcome is selected based on its significant impact on or association with neonatal health.

| | Normal pregnancy outcome | Adverse pregnancy outcome |
|---|---|---|
| Maternal and Labour | | |
| Maternal age (years) | 31.0 (27.0-35.0) | 31.0 (27.0-35.0) |
| Maternal BMI (kg/m$^2$) | 26.1 (22.9-30.5) | 26.3 (23.0-30.7) |
| Gravidity | 1 (0-2) | 1 (0-2) |
| Parity | 0 (0-1) | 0 (0-1) |
| FHR trace duration (mins) | 60.0 (55.0-60.0) | 60.0 (60.0-60.0) |
| Gestational age at FHR trace | 34.5 (30.0-38.0) | 34.5 (30.0-38.0) |
| Labour onset | | |
| Induced | 280 (29.9%) | 158 (16.9%) |
| Spontaneous | 508 (54.2%) | 105 (11.2%) |
| No labour | 149 (15.9%) | 673 (71.8%) |
| Delivery method | | |
| Spontaneous Vertex | 589 (62.9%) | 0 (0.0%) |
| Forceps | 153 (16.3%) | 0 (0.0%) |
| Ventouse | 46 (4.9%) | 0 (0.0%) |
| Breech | 0 (0.0%) | 0 (0.0%) |
| Emergency CS | 0 (0.0%) | 264 (28.2%) |
| Elective CS | 149 (15.9%) | 673 (71.8%) |
| Newborn | | |
| Gestational age at delivery | 39.0 (38.0-40.0) | 35.0 (30.0-38.0) |
| Male | 510 (54.3%) | 513 (54.6%) |
| Female | 430 (45.7%) | 427 (45.4%) |
| Apgar | | |
| 1 minute | 10.0 (9.0-10.0) | 3.0 (2.0-5.0) |
| 5 minutes | 10.0 (10.0-10.0) | 8.0 (6.0-10.0) |
| 10 minutes | 10.0 (10.0-10.0) | 10.0 (8.0-10.0) |
| Arterial pH | 7.25 (7.20-7.30) | 7.21 (7.10-7.27) |
| Venous pH | 7.33 (7.28-7.36) | 7.26 (7.16-7.32) |
| Arterial base excess | 5.45 (3.40-8.20) | 5.50 (3.40-10.30) |
| Venous base excess | 4.50 (2.50-6.10) | 5.00 (2.30-8.45) |

**Supplementary Table 2: Maternal, labour, and newborn characteristics stratified by pregnancy outcome.** Data are presented as median (interquartile range) for continuous variables and number (percentage) for categorical variables. Abbreviations: BMI, body mass index; FHR, fetal heart rate; CS, caesarean section.

| FHR Pattern | NPO | APO | Effect size | p-value |
|---|---|---|---|---|
| BHR | 142.0 (136.0–149.0) | 143.0 (136.2–150.0) | -0.09 | 0.14 |
| Accelerations | 8.0 (4.0–13.0) | 2.0 (0.0–5.0) | 0.93 | *** |
| Decelerations | 1.0 (0.0–2.0) | 1.0 (0.0–2.0) | -0.22 | *** |
| LDM | 11.0 (8.0–25.0) | 20.0 (10.0–41.0) | -0.17 | *** |
| STV | 7.7 (6.3–9.5) | 4.9 (3.8–6.6) | 1.01 | *** |
| High Variation | 0.3 (0.1–0.6) | 0.0 (0.0–0.2) | 0.91 | *** |
| Low Variation | 0.1 (0.0–0.3) | 0.5 (0.2–0.8) | -1.11 | *** |

Supplementary Table 3: Comparison of antepartum preterm fetal heart rate (FHR) patterns between normal pregnancy outcomes (NPO) and adverse pregnancy outcomes (APO). Data are presented as median (interquartile range). Effect size and p-values indicate the magnitude and significance of differences. Abbreviations: LDM, largest deceleration magnitude; High Variation and Low Variation represent the proportion of the total length of the FHR trace.

| FHR Pattern | NPO | APO | Effect size | p-value |
| --- | --- | --- | --- | --- |
| BHR | 136.0 (129.0–144.0) | 141.0 (132.0–149.0) | -0.38 | *** |
| Accelerations | 11.0 (6.0–17.0) | 6.0 (2.0–10.0) | 0.73 | *** |
| Decelerations | 0.0 (0.0–1.0) | 1.0 (0.0–1.0) | 0.01 | 0.92 |
| LDM | 10.0 (8.0–18.0) | 12.0 (8.0–25.0) | -0.03 | * |
| STV | 8.4 (6.7–10.6) | 6.4 (5.0–8.6) | 0.6 | *** |
| High Variation | 0.4 (0.2–0.6) | 0.1 (0.0–0.4) | 0.63 | *** |
| Low Variation | 0.2 (0.0–0.4) | 0.3 (0.1–0.6) | -0.56 | *** |

**Supplementary Table 4: Comparison of antepartum term fetal heart rate (FHR) patterns between normal pregnancy outcomes (NPO) and adverse pregnancy outcomes (APO).** Data are presented as median (interquartile range). Effect size and p-values indicate the magnitude and significance of differences. Abbreviations: LDM, largest deceleration magnitude; High Variation and Low Variation represent the proportion of the total length of the FHR trace.

| | Pattern | NPO | APO | Effect size | p-value |
|---|---|---|---|---|---|
| Acidaemia | BHR | 141.0 (133.0–146.8) | 145.5 (139.0–152.0) | -0.52 | *** |
| | Accelerations | 9.5 (6.0–16.0) | 1.0 (0.0–4.0) | 1.58 | *** |
| | Decelerations | 1.0 (0.0–1.0) | 1.0 (0.0–2.0) | -0.35 | * |
| | LDM | 10.0 (8.0–19.8) | 18.5 (10.0–39.0) | -0.4 | *** |
| | STV | 8.1 (6.6–10.0) | 4.4 (3.4–6.0) | 1.4 | *** |
| | High Variation | 0.4 (0.2–0.6) | 0.0 (0.0–0.1) | 1.38 | *** |
| | Low Variation | 0.1 (0.0–0.3) | 0.6 (0.3–0.8) | -1.47 | *** |
| Asphyxia | BHR | 138.0 (131.5–146.0) | 146.0 (140.0–153.0) | -0.73 | *** |
| | Accel | 9.0 (6.0–17.0) | 1.0 (0.0–4.0) | 1.61 | *** |
| | Decelerations | 0.0 (0.0–1.0) | 1.0 (0.0–2.0) | -0.38 | 0.09 |
| | LDM | 15.5 (9.8–25.0) | 21.0 (9.0–37.5) | -0.15 | 0.28 |
| | STV | 8.7 (6.9–10.3) | 4.4 (3.4–6.0) | 1.58 | *** |
| | High Variation | 0.4 (0.1–0.6) | 0.0 (0.0–0.1) | 1.28 | *** |
| | Low Variation | 0.1 (0.0–0.3) | 0.5 (0.3–0.8) | -1.38 | *** |
| Stillbirth | BHR | 143.0 (136.2–148.0) | 143.5 (138.2–153.0) | -0.3 | 0.11 |
| | Accelerations | 8.5 (4.0–14.0) | 2.0 (0.0–5.0) | 1.2 | *** |
| | Decelerations | 1.0 (0.0–2.0) | 1.0 (0.0–2.0) | -0.08 | 0.99 |
| | LDM | 10.0 (7.0–15.0) | 18.0 (9.8–35.2) | -0.62 | ** |
| | STV | 7.3 (6.0–9.2) | 4.8 (3.6–6.2) | 1.23 | *** |
| | High Variation | 0.2 (0.1–0.6) | 0.0 (0.0–0.1) | 1.09 | *** |
| | Low Variation | 0.1 (0.0–0.3) | 0.5 (0.3–0.8) | -1.19 | *** |
| HIE | BHR | 133.0 (126.5–139.0) | 141.0 (138.5–147.5) | -1.04 | *** |
| | Accelerations | 13.0 (5.5–20.0) | 3.0 (0.0–6.0) | 1.33 | *** |
| | Decelerations | 0.0 (0.0–0.5) | 0.0 (0.0–1.0) | -0.4 | 0.17 |
| | LDM | 7.5 (7.0–9.2) | 9.0 (7.0–12.0) | -0.56 | 0.21 |
| | STV | 9.8 (8.5–11.8) | 5.8 (3.3–6.9) | 1.83 | *** |
| | High Variation | 0.6 (0.3–0.8) | 0.0 (0.0–0.2) | 1.46 | *** |
| | Low Variation | 0.1 (0.0–0.3) | 0.6 (0.3–0.8) | -1.37 | *** |
| Low Apgar | BHR | 140.0 (134.0–148.0) | 142.0 (134.0–150.0) | -0.16 | * |
| | Accelerations | 9.0 (4.0–15.0) | 2.0 (0.0–8.0) | 0.77 | *** |
| | Decelerations | 1.0 (0.0–2.0) | 1.0 (0.0–2.0) | -0.07 | 0.13 |
| | LDM | 11.0 (8.0–25.0) | 16.0 (9.0–37.2) | -0.08 | *** |
| | STV | 7.8 (6.4–9.8) | 5.5 (4.2–7.5) | 0.8 | *** |
| | High Variation | 0.3 (0.1–0.6) | 0.1 (0.0–0.3) | 0.73 | *** |
| | Low Variation | 0.1 (0.0–0.4) | 0.5 (0.2–0.7) | -0.9 | *** |
| Extended NU | BHR | 137.0 (131.0–145.2) | 142.0 (134.8–150.0) | -0.38 | *** |
| | Accelerations | 11.5 (7.0–17.2) | 6.0 (2.0–9.0) | 0.87 | *** |
| | Decelerations | 0.0 (0.0–1.2) | 1.0 (0.0–2.0) | -0.2 | * |
| | LDM | 10.0 (7.0–19.0) | 13.0 (9.0–29.5) | -0.27 | * |
| | STV | 8.4 (6.7–10.8) | 6.5 (5.0–8.2) | 0.67 | *** |
| | High Variation | 0.4 (0.2–0.7) | 0.1 (0.0–0.3) | 0.76 | *** |
| | Low Variation | 0.2 (0.0–0.4) | 0.3 (0.1–0.6) | -0.56 | *** |
| Resuscitation | BHR | 141.0 (132.0–147.0) | 143.0 (138.0–148.0) | -0.46 | 0.05 |
| | Accelerations | 9.0 (3.0–17.0) | 2.0 (0.0–7.0) | 0.95 | *** |
| | Decelerations | 1.0 (0.0–2.0) | 1.0 (0.0–1.0) | 0.03 | 0.51 |
| | LDM | 14.0 (8.0–26.5) | 12.0 (9.0–48.0) | 0.11 | 0.52 |
| | STV | 8.7 (6.7–11.4) | 5.6 (4.2–7.4) | 1.15 | *** |
| | High Variation | 0.4 (0.1–0.7) | 0.0 (0.0–0.3) | 0.99 | *** |
| | Low Variation | 0.0 (0.0–0.3) | 0.5 (0.1–0.8) | -1.04 | *** |

**Supplementary Table 5: Comparison of fetal heart rate (FHR) patterns across different adverse pregnancy outcomes (acidaemia, asphyxia, stillbirth, hypoxic ischaemic encephalopathy [HIE], low Apgar, Extended neonatal care unit admission (NCU), resuscitation) versus normal pregnancy outcomes (NPO).** Data are presented as median (interquartile range). Effect size and p-values indicate the magnitude and significance of differences. Abbreviations: NU, neonatal unit; LDM, largest deceleration magnitude; Group sizes: Acidaemia = 150, Asphyxia = 67, Stillbirth = 85, HIE = 31, Low Apgar = 695, Extended NCU = 164, Resuscitation = 65.
